# Post-treatment downregulation of type III interferons in patients with acute Brucellosis

**DOI:** 10.1101/2021.04.17.21255534

**Authors:** Mehran Shokri, Oreinab Ghaffari Khonakdar, Mousa Mohammadnia-Afrouzi, Mahmoud Sadeghi-Haddad-Zavareh, Amirhossein Hasanpour, Mohammad Barary, Soheil Ebrahimpour

## Abstract

There is a limited number of clinical studies on interferon (IFN) levels in human brucellosis. The novel group of interferons, type III interferons, consists of four IFN-λ (lambda) molecules called IFN-λ1 or Interleukin-29 (IL-29), IFN-λ2 or IL-28A, IFN-λ3 or IL-28B, and IFN-λ4, are not fully known. This study is one of the first studies of IL-28A and IL-29 levels in Brucellosis cases at the end of their treatment course. A total of 33 acute Brucellosis patients were included in this study. We considered changes in the levels of IL-28A and IL-29 in cases with acute brucellosis before and after treatment with standard therapy that referred to the Ayatollah Rohani Hospital in Babol, Northern Iran. Serum IL-29 and IL-28A (acute form: 56.4 ± 30.32 and 48.73 ± 27.72, respectively, and post-treatment: 40.15 ± 20.30 and 38.79 ± 22.66, respectively) levels were elevated significantly in acute brucellosis than after treatment (p ≤ 0.05). These findings indicate that considering biomarker levels in Brucellosis patients may be indicative of the chronicity of infection. In conclusion, we suggest that IL-29 and IL-28A levels may be valuable biomarkers for follow-up patients with brucellosis.

## Introduction

Brucellosis is a zoonotic bacterial disease caused by one of the various species of the *Brucella spp*. (1–3). Although approximately half-million new Brucellosis cases are reported worldwide, the actual incidence rate has been much more significant (4,5). Even though the gold standard for diagnosing this disease is leukocyte culture, this test has a high false-negative rate. Moreover, its cost and a 10-day delay before confirmation restrict its use as a standard diagnostic test in acute Brucellosis (6). As a result, clinicians chiefly rely on other laboratory tests to evaluate patients with brucellosis, such as agglutination test, white blood cell (WBC) counts, platelet (PLT) counts, liver function tests, erythrocyte sedimentation rate (ESR), and C-reactive protein (CRP). Nevertheless, the diagnosis of brucellosis remains a challenge in most cases (6).

Although rare, Brucellosis infection has a chronic disease course that may continue to trouble patients for years. No approved human anti-Brucellosis vaccine is currently available (7). So far, the Brucellosis studies have been focused primarily on epidemiological investigations, and the immune response against these bacteria was somehow neglected (8). After the entry of this pathogen, several immunogenic changes are prominent in the host body. For example, interferon-gamma (INF-γ) and tumor necrosis factor-alpha (TNF-α) levels are elevated as naïve T cells differentiate to CD4^+^ helper T cells type 1 (Th_1_) and interleukin-4 (IL-4) levels are increased as naïve T cells give rise to CD4^+^ helper T cells type 2 (Th_2_) (6).

Moreover, transforming growth factor-beta (TGF-β) levels are also raised due to the surge in regulatory T cells (Tregs) population (6). Several studies have shown that interferon-gamma serum concentration may be recognized as an essential factor in chronic Brucellosis (8). It is vital to note the association between other types of cytokines, such as type III interferons or interferon-lambda (IFN-λ) and the clinical course of brucellosis, including response to treatment, is not yet fully understood. Nevertheless, some studies recognized bacterial pathogens might activate IFN-λ1 or Interleukin-29 (IL-29), IFN-λ2 or IL-28A, IFN-λ3, or IL-28B (9,10).

Therefore, this study was conducted to evaluate the levels of IL-28A and IL-29 in patients with brucellosis, both pre-treatment, and post-treatment.

## Materials and Methods

### Patients

In this case-control study at the Ayatollah Rohani Hospital in Babol, northern Iran, 33 pre-treatment and post-treatment acute Brucellosis patients were included. Inclusions criteria were defined as receiving a clinical diagnosis of acute brucellosis (clinical presentation time: acute form (≤ 2 months) based on the symptoms, compatible clinical findings, standard tube agglutination (STA) test titer ≥ 1:160, and in the presence of 2-mercaptoethanol (2ME) agglutination ≥ 1:80. The control group consisted of the same 33 patients who had undergone a complete course of treatment, STA, and 2ME tests and did not meet the above clinical and laboratory criteria. The exclusion criteria were pregnancy, age < 18 years, and other chronic infectious or immunodeficiency diseases. All patients received gentamicin 5 mg/kg/day for one week plus doxycycline 100 mg BID for 45 days. Informed consent was obtained from all study participants.

### Determination of cytokine levels

Blood samples were collected in an ethylenediamine tetra-acetic acid (EDTA) containing tube. One blood sample (5 ml) was obtained from all patients before and after treatment. The blood samples were centrifuged at 400 g for 35 minutes, and collected sera were stored at -80°C for further analysis. IL-28A and IL-29 levels in all serum samples were measured using a commercial enzyme-linked immunosorbent assay (ELISA) kit (eBioscience, San Diego, CA) according to the manufacture’s guidelines.

### Statistical analysis

All data are expressed as mean ± standard deviation (SD). Statistical analysis was performed using the SPSS software version 16.0 (IBM, Chicago, IL, USA). The difference between groups was analyzed using the paired t-test, and the relationship between variables was evaluated using Spearman’s rank correlation test. A p-value of ≤ 0.05 was defined as statistically significant.

## Results

Of 33 included patients, 22 (66.6%) were males, and 11 (33.4%) were females. The range of patients’ age was 49.21 ± 17.70 years.

Serum levels of IL-28A and IL-29 in the study groups (before and after the treatment) are illustrated in Figure 1. Serum IL-28A levels were 48.73 ± 27.72 and 38.79 ± 22.66 pg/mL in pre-and post-treatment, respectively (p = 0.038) (Figure 1). Also, IL-29 serum levels were 56.45 ± 30.32 and 40.15 ± 20.30 pg/mL in pre-and post-treatment, respectively (p = 0.026) (Figure 1). Also, it is noteworthy that IL-29 levels both before and after treatment were more than IL-28A levels.

**Figure 1.**
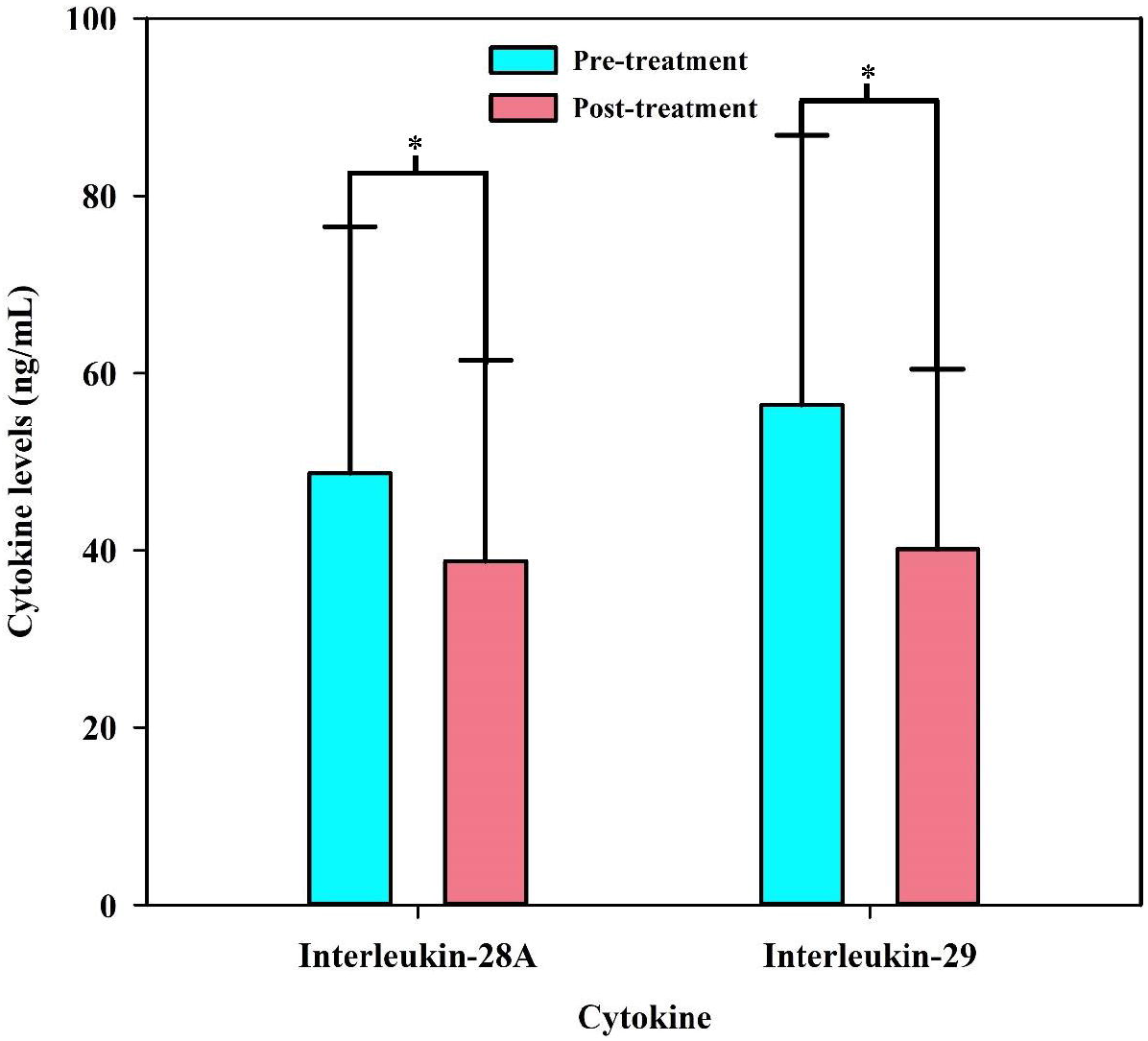
Serum levels of interleukin-28A and interleukin-29 among patients with brucellosis before and after treatment. All data are presented as mean ± SD. ^*^ Indicates p < 0.05.

## Discussion

In this case-control study, the IL-28A and IL-29 levels of 33 patients with confirmed acute brucellosis were measured both pre-and post-treatment. It was observed that the levels of these biomarkers were significantly decreased after the eradication of the disease.

Several studies revealed the crucial role of type III IFNs for resistance to viral infections, principally by induction of the antiviral state. Furthermore, some studies revealed that type III IFNs play a significant role in inhibiting virus replication by mediating and expressing interferon-regulated genes (IRGs) (11). Subsequent studies have also shown an inhibiting role of interferon in replicating Zika virus (ZIKV), influenza A and B viruses, coronavirus, and respiratory syncytial virus (RSV) (12,13). Ank et al. concluded that interferon-lambda plays a crucial role in the innate immune response through activating the macrophages and dendritic cells against human Herpesvirus type-1 (HHV-1) (14). Another study demonstrated Dengue virus (DENV) replication inhibition through interferon-regulating gene expression (15).

The role of IL-29 in bacterial and parasitic infections and its increased expression in these diseases has been proven before. Also, the critical role of interferon-lambda has been highlighted in the acquired immune response in previous studies. This cytokine initiates the received immune response through its effect on antigen-presenting cells. Moreover, type III IFNs suppresses the immune response through regulatory T cells (Tregs), promoting the acquired immune response (16). As shown in previous studies, the defense of the human immune system against brucellosis is dependent on cellular immunity, which mainly affects antigen-presenting cells (APC), such as macrophages, dendritic cells, and CD4^+^ and CD8^+^ T cells. Other defense cells, such as natural killer (NK) cells and other T lymphocytes, also play a crucial role in cellular immunity against *Brucella* (17). As a result, the Th_1_-dependent immune response is dominant at the onset of *Brucella* infection and is critical in eradicating the disease. Therefore, this issue fully justifies the current study results, particularly the higher levels of IL-29 in patients with acute brucellosis before treatment than after receiving the standard treatment (17). Figure 2 illustrates a summary of the immune reactions of the host’s body and the vital role of different cytokines, such as IFN-γ, TNF-α, and IFN-λ, in the eradication of *Brucella spp*.

**Figure 2.**
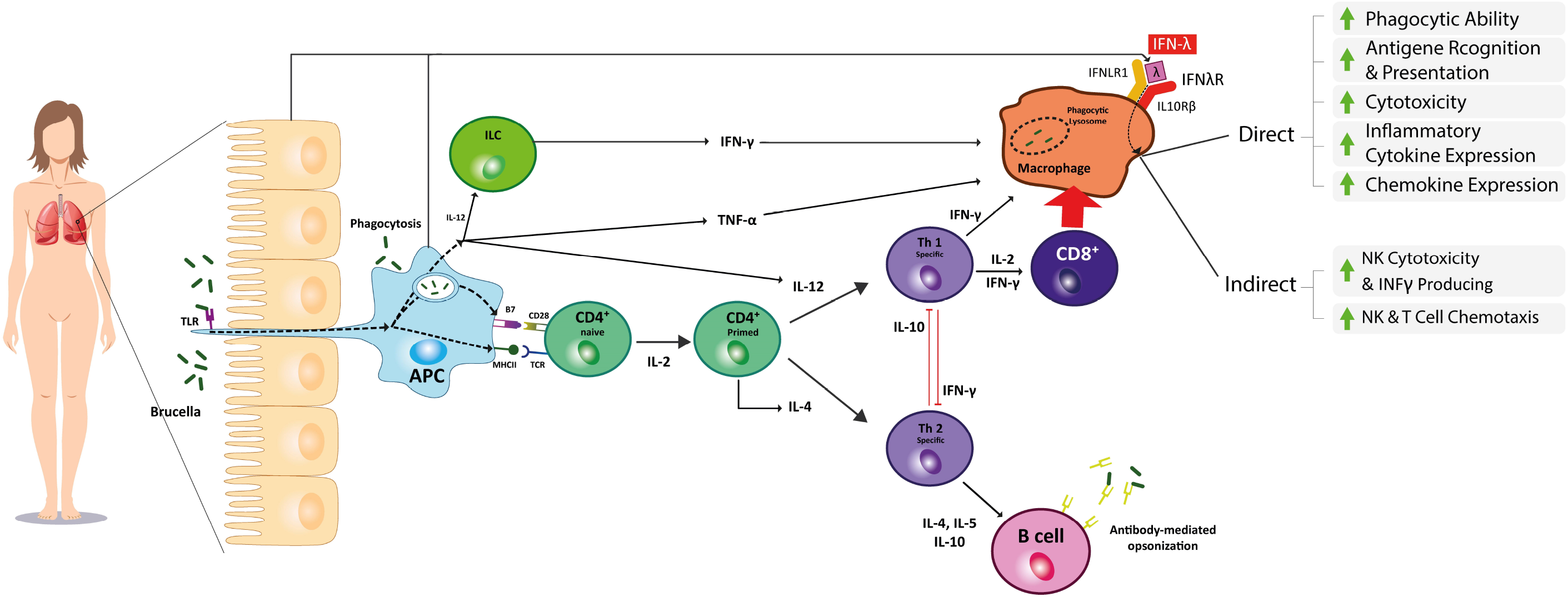
Interactions of *Brucella spp*. With the immune system. After the activation of antigen-presenting cells (APCs) with *Brucella* antigen via toll-like receptor (TLR) signaling pathway, a cascade of events leads to the priming of CD4^+^ T cells to helper T cells type 1 (Th_1_) and type 2 (Th_2_). Th_1_ cells secrete various cytokines, such as TNF-α, and IFN-γ, which activate and enhance the anti-brucella mechanisms of macrophages and activate CD8^+^ T cells, which boost the immune responses of macrophages even further. Moreover, APCs can trigger Th_2_ activation, which switches on B lymphocytes and the humoral immunity, facilitating the opsonization and faster eradication of the pathogen from the host’s body. It is noteworthy that Th_1_ and Th_2_ cells can inhibit either pathway via secreting cytokines, such as IFN-γ, and IL-10, respectively. Furthermore, macrophages can be stimulated by secreting another cytokine called type III interferons or interferon-λ by APCs or epithelial cells. These activated macrophages exert their immunomodulatory effects through two different pathways: direct and indirect. In the direct pathway, chemokine and inflammatory cytokine expression, antigen recognition and presentation, and macrophages’ cytotoxicity are elevated. Through the indirect pathway, these cells can enhance natural killer (NK) and T cells chemotaxis, NK cells cytotoxicity, and elevate the production and release rate of IFN-γ, which in turn, via activating the Th_1_ pathway, helps better and faster eradication of *Brucella spp*. Abbreviations: TLR, Toll-like receptor; ILC, Innate lymphocyte cells; IL-12, Interleukin-12; APC, Antigen-presenting cells; B7, Cluster of differentiation 80/86; MHC II, Major histocompatibility complex type 2; CD28, Cluster of differentiation 28; TCR, T cell receptor; IFN-γ, Interferon-gamma, TNF-α, Tumor necrosis factor-alpha; IL-4, Interleukin-4; Th_1_, Helper T cell type 1; Th_2_, Helper T cell type 2; IL-2, Interleukin-2; IL-10, Interleukin-10; IL-5, Interleukin-5; IFN-λ, Interferon-λ; IFNLR1, Interferon-lambda receptor 1; IL10Rβ, Interleukin-10 receptor beta; IFNλR, Interferon-lambda receptor; NK cell, Natural killer cell

In the present study, we showed a significant decrease in IL28-A and IL-29 levels after treatment with the standard antibiotic regimen, i.e., gentamicin 5 mg/kg QD for one week plus doxycycline 100 mg BID for 45 days. Regarding the IL-28A levels, it seems that cross-linking of IL-28A with type I interferons (IFNs) and subsequent innate and acquired immune responses against brucellosis may be the cause. For IL-29, this reduction can be attributed to its vital role in acquired immunity, eradicating brucellosis using antibiotic treatment, and reduced inflammation (17). IFNs initiate an innate immune response after contact with pathogens. The immune response and immune mediators and the subsequent inflammation are expected to decline following the control and eradication of infection. It is important to note that some standard diagnostic tests, such as STA, Coombs Wright, and 2ME, may remain positive even at high titers for up to two years after treatment (18,19). Thus, such tests’ application was not justified for follow-up patients. Practically, most patients are generally followed up with their symptoms (20).

The current study results revealed a significant reduction in serum IL-28A and IL-29 levels after treatment, making these biomarkers a valuable indicator for monitoring the patients. While the effects of IL-28A on the acquired immunity are not significant, its functions are chiefly exerted via innate immunity, with no effect on increasing or decreasing immunoglobulins (Ig) (20). On the other hand, some previous studies have shown a positive correlation of IL-29 with serological tests that indicate its impact on acquired immunity and Ig production. However, it is essential to note that cellular immunity plays a more crucial role in eliminating Brucellosis (20).

The primary limitation of this was our small sample size. It is recommended to reperform such studies in a larger sample size. Also, another limitation of the current study was the sole evaluation of interferon-lambda. It is suggested that future researchers assess the levels of other vital cytokines, such as interferon-alpha (IFN-α) and -gamma (IFN-γ), tumor necrosis factor-alpha (TNF-α), transforming growth factor-beta (TGF-β), interleukin-2 (IL-2), and interleukin-12 (IL-12).

In conclusion, this study’s findings confirmed previous studies on bacterial infections and validated the pivotal role of IFN-λ during acute brucellosis via strengthening innate and promoting acquired immunities. Therefore, the significant reduction in serum levels of IL-28A and IL-29 in patients with brucellosis after a standard treatment regimen may promise the emergence of valuable biomarkers in patient follow-up.

## Data Availability

The data that support the findings of this study are available from the corresponding author upon reasonable request.

## Acknowledgments

The authors thank the Department of Infectious diseases of Babol University of Medical Sciences.

## Notes

### Competing Interest Statement

The authors have declared no competing interest.

### Funding Statement

This study was fully supported by the vice-chancellor for research and technology of Babol University of Medical Sciences.

### Author Declarations

This study protocol was approved by the ethics committee of Babol University of Medical Sciences (IR.MUBABOL.HRI.REC.1397.212).

